# Pulse-Controlled Amplification – a new powerful tool for front-line diagnostics

**DOI:** 10.1101/2020.07.01.20144097

**Authors:** Katharina Müller, Sarah Daßen, Scott Holowachuk, Katrin Zwirglmaier, Joachim Stehr, Federico Buersgens, Lars Ullerich, Kilian Stoecker

## Abstract

Molecular diagnostics has become essential in the identification of many infectious diseases, and the detection of nucleic acids often serves as the gold standard technique for most infectious agents. However, established techniques like polymerase chain reaction (PCR) are time-consuming laboratory-bound techniques. Here we present an alternative method for the rapid identification of infectious agents using pulse-controlled amplification (PCA). PCA is a next generation nucleic acid amplification technology that uses rapid energy pulses to heat microcyclers (micro-scale metal heating elements embedded directly in the amplification reaction) for a few microseconds, thus only heating a small fraction of the reaction volume. The heated microcyclers cool off nearly instantaneously, resulting in ultra-fast heating and cooling cycles during which classic amplification of a target sequence takes place. This reduces the overall amplification time by a factor of up to 10, enabling a sample-to-result workflow in just 15 minutes, while running on a small and portable prototype device. We could demonstrate the efficacy of this technology in two assays, one for a nosocomial context targeting MecA conferred antibiotic resistance, and one for a biothreat scenario targeting *Yersinia pestis*. The observed limits of detection were 10 copies per reaction (purified DNA) for MecA in a methicillin resistant *Staphylococcus aureus* and 434 copies per reaction (purified DNA) or 9.8 cells per reaction (crude sample) of *Yersinia pestis*. Thus, PCA offers a decentralization of molecular diagnostics and is applicable whenever rapid, on-site detection of infectious agents is needed.

## INTRODUCTION

Infectious diseases caused by pathogenic microorganisms such as bacteria, viruses, fungi or parasites, are still among the top ten causes of death worldwide (1).Rapid and specific diagnostics are an essential pillar of every outbreak-response and can prevent epidemics or even pandemics. One infamous example is pneumonic plague caused by *Yersinia pestis*, a category A bioterrorism agent, (2) which can be fatal if not treated within 24 hours after the onset of symptoms. Furthermore, modern threats of bioterrorism have the potential to cause mass casualty incidents of serious infectious diseases, and therefore require quick and reliable diagnostic tests that can be performed on-site rather than in a centralized laboratory (3). In a clinical setting rapid on-site screening of patients with a suspected MRSA-infection is an essential for treatment and prevention of nosocomial spread. Thus, in modern diagnostics, rapid diagnostic tests (RDTs) are indispensable assets, but have to live up to high expectations: they need to be portable and easy-to-use, while at the same time delivering results with high sensitivity and specificity in little time, in order to facilitate a reliable diagnosis for adequate therapy.

Lateral flow assays (LFAs) are a common example of RDTs and come in several different formats. Principally, they all use immunochromatography to detect the presence or absence of a specific target analyte in a liquid sample without the need for intensive sample preparation and additional equipment. They are small and portable devices that deliver rapid (i.e. 15-20 minutes) results and thereby moving diagnostic testing away from centralized laboratories and closer to the patient or even directly into the field. Thus, they have become an indispensable technique in point of care (POC) testing and are used by first responders worldwide (4,5). However, data on sensitivity and specificity of LFAs is limited and generally points to a high detection limit (6). At the same time serological detection of antigens is known to be challenging as antibodies often show cross-reactivity with other non-target antigens, leading to false positive results (7). Furthermore, sensitivity of detection methods based on antigen recognition is dependent on the permanent presence and sufficient concentration of the specific antigen. LFAs for the detection of *Y. pestis*, for example, use the F1 capsular protein. However, the F1 gene is temperature regulated and only expressed at ≥33 °C, which can lead to false negative results (8).

In contrast to LFAs, polymerase chain reaction (PCR) a highly sensitive method with great specificity and reproducibility and many gold-standard protocols have been established for a broad range of biological agents. During the last decade, a variety of PCR-based techniques have been developed to further improve diagnostic capabilities, including real-time quantitative PCR (qPCR), multiplex PCR and digital droplet PCR (ddPCR) (9). Nevertheless, PCR remains a time-consuming and laboratory bound technique, usually requiring DNA extraction prior to amplification and large, heavy, and power-consuming thermal cyclers. Several other nucleic acid amplification methods are available which have the potential to become point of care detection and diagnostics tools, mostly relying on isothermal amplification methods such as Loop-mediated isothermal amplification (LAMP). However, they often require multiple primers, highly aggravating detection of groups of pathogens with highly divergent sequences, such as Filoviridae (10). Furthermore, the required large number of individual primers makes these assays especially prone to primer-dimer formation, which in some cases even lead to false positive results (11).

To address these different limitations that existing methods present, we introduce an ultra-fast, yet sensitive and specific method for the nucleic-acid based detection of infectious agents, i.e. pulse-controlled amplification (PCA). We demonstrate a simple and fast workflow for nucleic acid detection using PCA. To demonstrate the sensitivity of the PCA approach we used an assay for the detection of mecA, a gene conferring antibiotic resistance to many bacteria but most prominently to methicillin resistant staphylococcus aureus (MRSA). Moreover, we provide a proof of concept for the detection of *Y. pestis* under both standard lab-conditions (purified DNA) and in field use (crude sample).

## MATERIAL AND METHODS

### Pulse-controlled amplification

Similar to PCR, pulse-controlled amplification (PCA) relies on the exponential amplification of a specific nucleic acid target fragment for subsequent detection. Amplification is achieved through the binding of target-specific, complementary oligonucleotide primers to template DNA, followed by primer extension by a DNA polymerase enzyme. PCA also relies on thermal cycling, however, instead of time-consuming alternating heating and cooling of the total reaction volume (“global heating”), rapid sub-millisecond voltage pulses are applied to an array of 75 gold-coated tungsten wires (15 µm diameter, 200 nm Au coating), causing ultra-fast heating within only a micrometer-sized liquid layer surrounding each wire (“local heating”). The remaining bulk of the reaction volume (more than 99%) is kept at the base temperature used for annealing and elongation. The approach of “local heating” denatures double stranded (ds) DNA only within the heated layer surrounding the wires, which makes it necessary for part of the reaction to be localized as well. This is achieved by attaching one of the primers to the micro-scale conductive metal structures (in this study gold-coated tungsten wires). The other primer remains free in solution, providing the kinetic advantages of a free reaction. As a result, the dsDNA denaturation step of the amplification reaction requires only a fraction of the energy usually required to thermocycle the total reaction volume. Local heating allows the wires to cool off after the voltage pulse that drives the denaturation step by thermal diffusion on a millisecond time scale. The bulk of the reaction volume serves as cooling reservoir for an entirely passive cooling process of the embedded wires, resulting in ultra-fast thermal cycles. This reduces the total time of the amplification process by a factor of up to ten compared to PCR, as hundreds of energy pulses can take place in a short amount of time. Like qPCR, amplification can be traced in real time using intercalating dyes or, as in our study, hydrolysis probes (Fig. 1). Currently, PCA is performed on a prototype instrument, the Pharos Micro (GNA Biosolutions, Martinsried, Germany) utilizing prototype disposable chips, which contain the amplification reactions (GNA Biosolutions, Martinsried, Germany). To optimize assays, different parameters of the run are adjustable, including base and lid temperature of the Pharos Micro [°C], heating time [µs], cycle time [s], number of cycles, and thermalizing time [s]. For primer design, the guidelines in table 1 should be followed. For successful PCA, it is critical to avoid primer dimers when designing primers, especially for the thiolated-primer used for functionalization of the wires.

**Tab 1.**
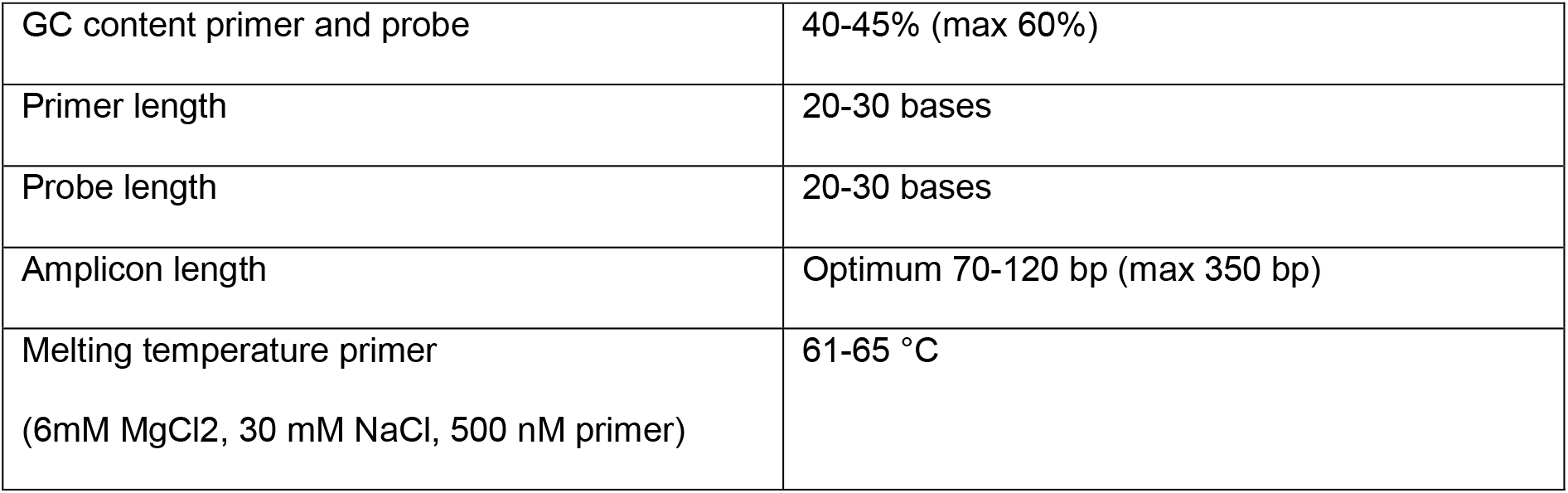
Guidelines for PCA primer and probe design.

**Fig 1:**
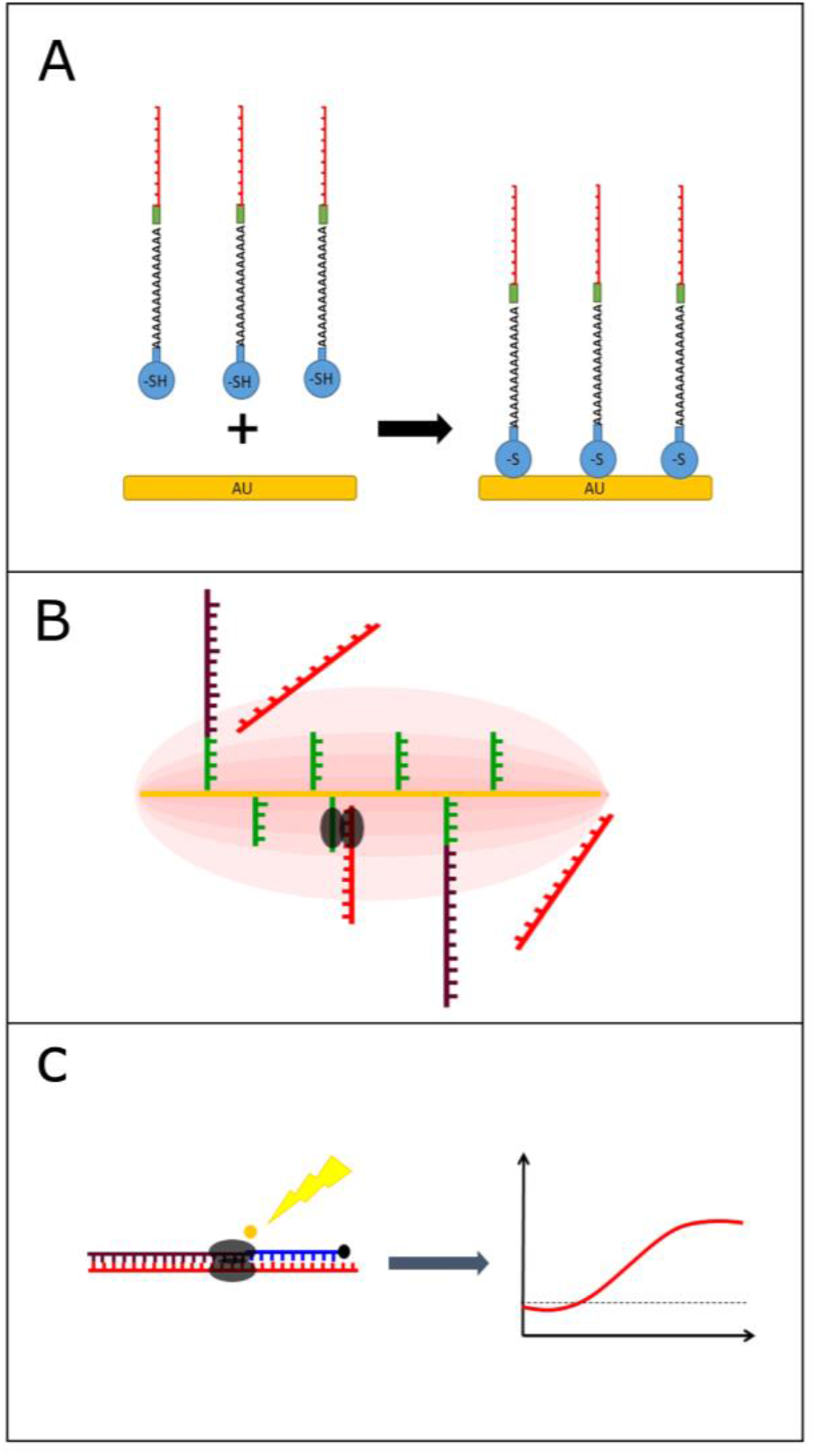
Schematic of the PCA process and the interaction of primers and gold coated wires. **(A)** (reduced) **Thiol-modification** was added to the 5’-end of the **primer** using a **poly-A-tail** and **Int Spacer 9**, allowing for a strong AU-S bond and immobilization of the primer to the gold-coated wires **(B)** PCA uses thermal cycling but instead of time-consuming alternating heating and cooling of the whole reaction mixture, rapid energy pulses are applied to the gold wires, causing ultra-fast heating and cooling of the reaction mixture surrounding the wires during which amplification can take place. **(C)** Amplification takes place in just 5-15 minutes and can be observed in real-time using hydrolysis probes

### Pharos Micro prototype and chip design

The Pharos Micro prototype (GNA Biosolutions, Martinsried, Germany) used in this study consists of a 3D-printed housing and lid with a size of 100mm x 175 mm x 110 mm (WxDxH) and weighs 900 g (Fig. 2). The instrument is equipped with light-emitting diodes and filters for real-time fluorescence detection and electronic control modules. Two conventional heating blocks are set to a constant temperature at the bottom and the top of the chip to maintain the reaction volume at a constant temperature of 65 °C for annealing and elongation. The Pharos Micro prototype is battery-powered for field use (using a commercially available power bank) or can be connected to a power supply (230 V) in the stationary lab. It is operated with a dedicated software (GNA Biosolutions, Martinsried, Germany) on a tablet (or laptop) connected to the instrument via USB. The prototype disposable test chips consist of Poly(methyl methacrylate) (PMMA) and have eight wells, with each one fitting 40-80 µl reaction volume (Fig. 2). All wells are equipped with 75 paralleled, gold-coated tungsten micro wires of 15 µm diameter resulting in a resistance of ≈ 500 mΩ. The wires allow for the electrical contact to the Pharos Micro prototype at both ends. Primers are coupled to the wires via a functionalization step. Chips are sealed with an adhesive tape after sample loading. More details on the prototype can be found in the SOMS material and methods.

**Fig 2:**
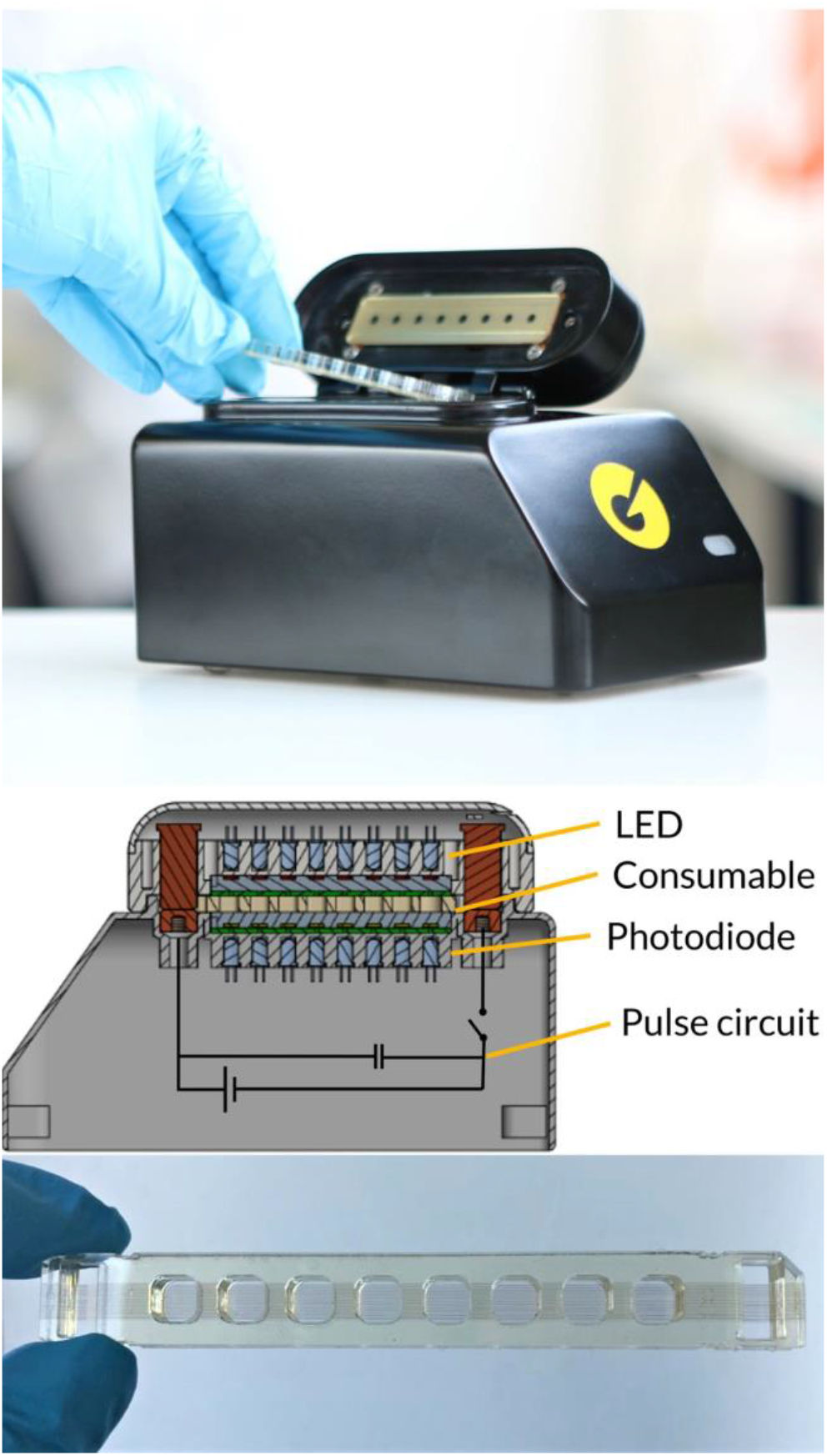
Pharos Micro prototype. Upper panel: The current model 8-well duplex manual sample-to- answer prototype system for research and assay development, using the PCA approach. **Middle panel:** Schematic view (right) showing the concept of the instrument: The disposable chip (yellow) is sandwiched between two heat blocks (grey) used to set the base temperature. Transmission fluorescence measurements (multicolour LEDs on top, photodiodes on the bottom) for hydrolysis probe chemistry as well as a schematic of the circuit to drive PCA, which basically only requires a capacitor and a fast switch (i.e. a MOSFET) to deliver the pulses necessary for localized heating. **Lower panel:** prototype chip. Each PMMA-chip has eight wells with 75 ultra-thin gold-coated tungsten micro wires running through the entire chip at the bottom of every well.

### Functionalization of Chips

Chips were functionalized with the (thiol-modified) forward primer. Primer was diluted in functionalization buffer 2 (GNA Biosolutions, Martinsried, Germany) to a final concentration of 500 nM. 50 µl of primer dilution was loaded into each well and incubated for 20 minutes at room temperature. Functionalization solution was removed and wells were washed five times with demineralized water. After removing the water, the dry chips were stored at 4 °C until further use. Chips were prepared fresh every morning.

#### mecA PCA assay

Primers and probe were designed based on alignment studies using BLAST and the sequence database of the National Centre for Biotechnology Information (NCBI). The primer pair and probe showing best results in pilot experiments (data not shown) were used and are shown in table 2. The forward primer was modified as depicted in table 2, including a thiol-modification, a poly-A part and an internal spacer. The mecA assay was performed using a freshly prepared mastermix. Optimal concentrations for all components were determined in pilot experiments (data not shown). Final concentrations and components are shown in table 3.

**Tab 2.**
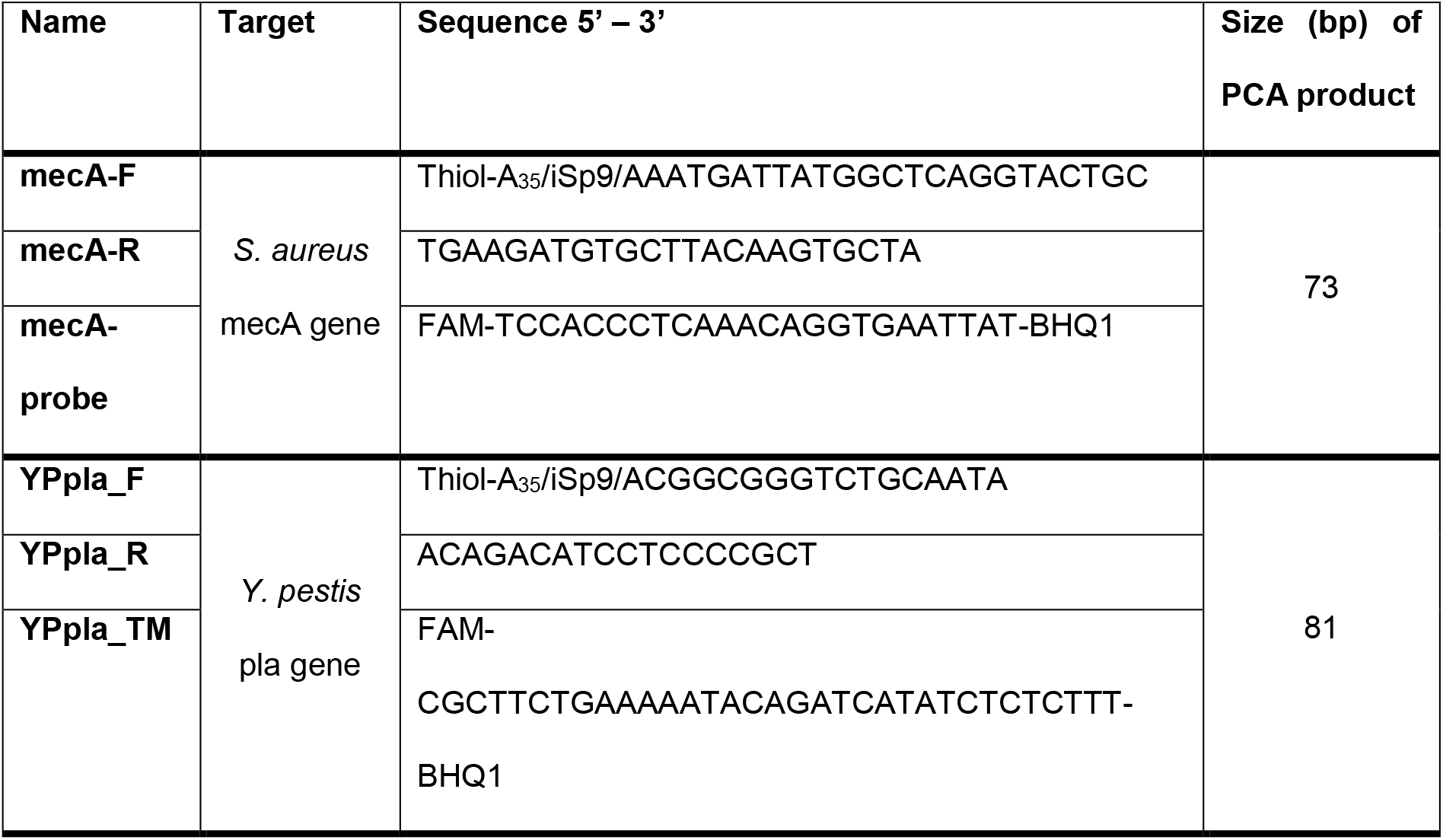
Primers and probes used in this study.

**Tab 3.**
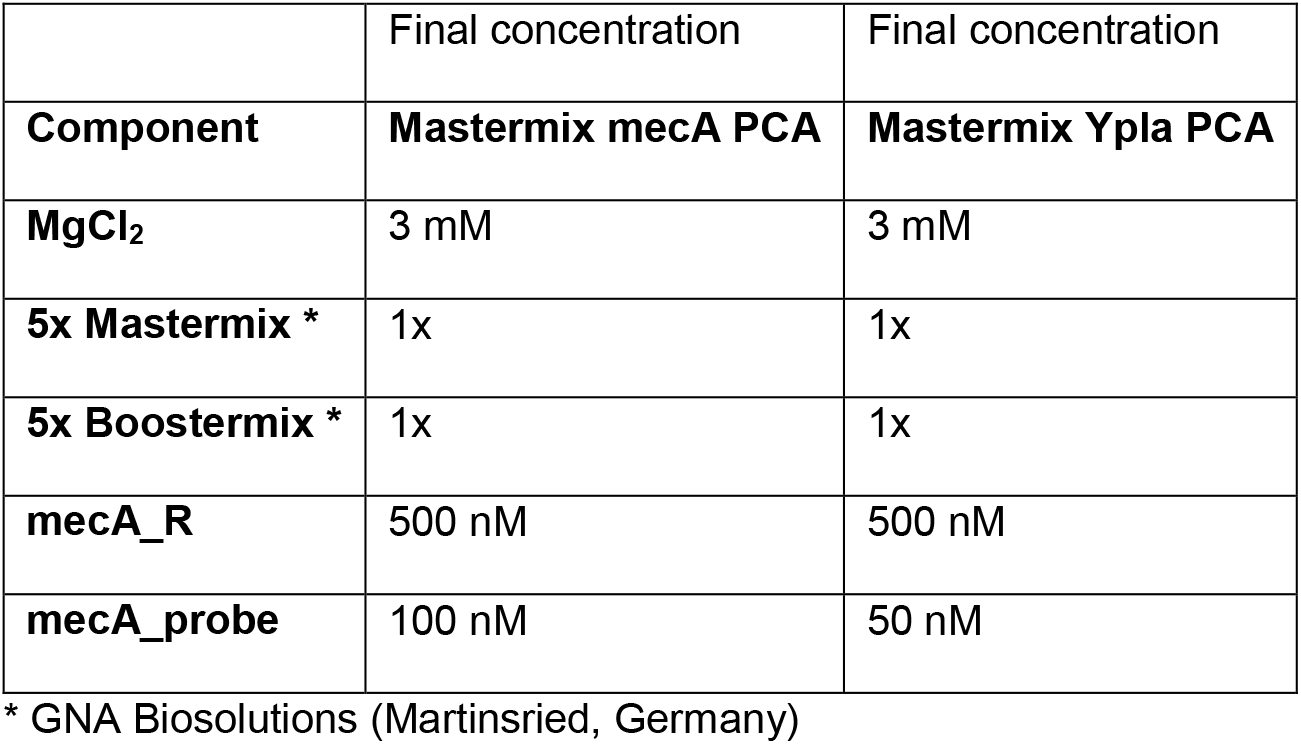
Optimized composition of the mastermixes for the mecA and Ypla PCA assays. The mastermix for the Ypla assay was freeze-dried.

**Tab 4.**
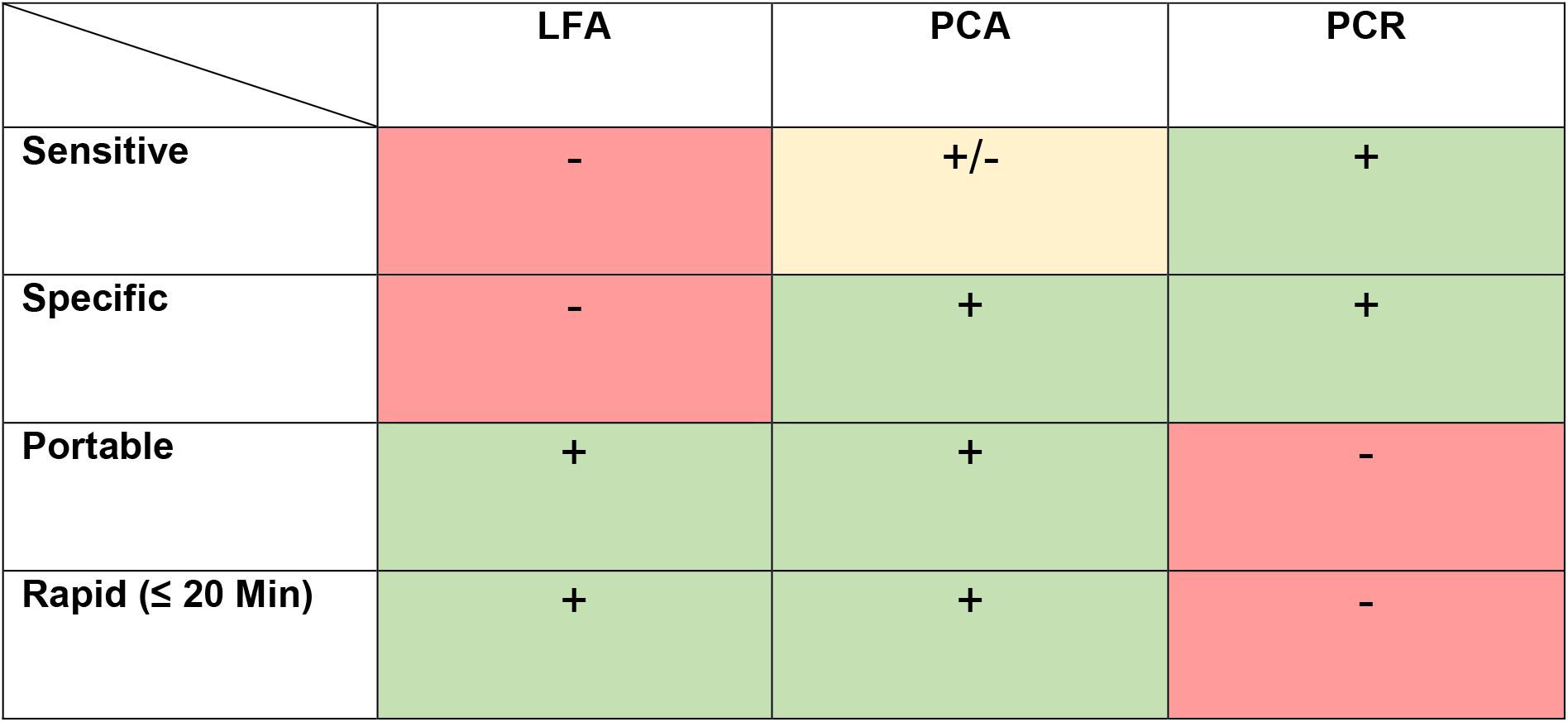
PCA is a cross-over technique of LFA and PCR. PCA is more sensitive and specific than LFAs while being equally rapid and portable. Compared to PCR it is slightly less sensitive but equally specific and only takes a fraction of the time required for PCR. In contrast to PCR it runs on a small portable platform.

For a PCA run the following parameters were used: 120 s thermalization, 800 cycles, 1.5 s cycle time, 71 °C lid temperature, 64 °C base temperature, 350 µs heating. Purified genomic DNA was denatured for 10 min at 99 °C (shorter times proved to work as well, data not shown) before serving as a template. 10 µl of the template was loaded into each well of the chip and 30 µl of the mastermix was added. Chips were sealed and placed in the Pharos Micro.

### Yersinia pestis pla PCA assay

*Y. pestis* pla gene specific primers and probes were designed based on alignment studies using BLAST and the sequence database of the National Centre for Biotechnology Information (NCBI). The primer pair and probe showing best results in pilot experiments (data not shown) were used for PCA experiments. Primers and probe were obtained from Ella Biotech (Martinsried, Germany). The forward primer was modified as depicted in table 2, including a Thiol-modification, a poly-A portion and an internal spacer. All reactions were performed using a freeze-dried mastermix (GNA Biosolutions, Martinsried, Germany), containing all necessary components. Optimal concentrations for all components were determined in pilot experiments (data not shown) and final concentrations are shown in table 3. For PCA, mastermix was dissolved in the appropriate amount of DNAse-free water to achieve a final reaction volume of 40 µl.

For a PCA run the following parameters were used: 10 s thermalization, 550 cycles, 1.5 s cycle time, 73 °C lid temperature, 66 °C base temperature, 350 µs heating.

For samples containing purified genomic DNA as template, lyophilized mastermix was dissolved in 316.8 µl DNAse-free water. 36 µl of mastermix was loaded into each well of the chip and 4 µl of template was added. For samples containing bacterial culture material, an initial thermal lysis and binding of the target DNA to the wires was performed prior to PCA: 60 µl/well of liquid culture material was loaded. Chips were sealed and placed in the Pharos Micro for 5 min (with 66 °C base temperature). After incubation, culture material was removed, lyophilized mastermix was dissolved in 352 µl DNAse free water and 40 µl was added directly to each well.

### Lateral Flow Assay

We used the miPROTECT Plague (Miprolab, Göttingen, Germany) lateral flow assay as the standard reference method for the detection of *Y. pestis* in liquid culture under field conditions. Tests were performed according to the protocol provided by the manufacturer and results were obtained after 20 minutes.

### Bacterial culture

*Y. pestis* strain EV76 was used as the model organism in this study. Liquid cultures were grown in LB– bouillon at 37 °C or at 28 °C to reduce expression of the F1 antigen. For comparability of experiments, *Y. pestis* cultures were diluted to a McFarland Standard 0.5 followed by serial 10-fold dilutions in LB-medium which were then used for experiments. Dilutions were prepared fresh every day.

For experiments with inactivated sample in the field exercise, a fixation of culture material was performed with 4% paraformaldehyde (PFA). For that, 1 ml of liquid culture was centrifuged at 14,000 rpm for 2 minutes. The supernatant was discarded and the cell pellet was resuspended in 1 ml PBS and centrifuged again. The supernatant was discarded, the cell pellet was resuspended in 250 µl PBS and 750 µl 4% PFA and fixated at 4 °C overnight. To remove PFA from the fixated sample, it was centrifuged at 14,000 rpm for 2 minutes, supernatant was discarded and the cell pellet was resuspended in 1 ml PBS and stored at 4 °C until further use.

### DNA preparation

For the mecA assay the *Staphylococcus aureus* DNA was commercially acquired at a concentration specified by the manufacturer (0801638DNA-10µg, ZeptoMetrix® Corporation, NY, USA). Serial 10-fold dilutions were prepared with TE buffer and stored at -20 °C until further use.

For DNA extraction, 1 ml of a liquid culture of *Y. pestis* with McFarland 0.5 was used. Purified DNA was prepared using the QIAamp DNA Mini Kit (Qiagen, Hilden, Germany) according to the manufacturer’s instructions. DNA was eluted in 50 µl of elution buffer and stored at -20 °C until further use. DNA concentration was determined using the Qubit dsDNA high-sensitivity (HS) quantification assay according to the manufacturer’s instructions. Measurement was performed using a Qubit 3.0 fluorometer (Invitrogen, Carlsbad, USA).

### Real-time quantitative PCR (qPCR)

qPCR was used as a reference method for the amplification of *Y. pestis* DNA. qPCR was performed as previously described by Riehm et. al. using the *pla* target region (12). In this study, sample volume was 5 µl and the assay was performed using a Rotor-Gene Q 2plex (Qiagen, Hilden, Germany).

### Digital droplet PCR

Since we used also whole cells as targets for the *Y. pestis* assay, we used digital droplet PCR (ddPCR) to determine accurate *Y. pestis* copy numbers, for both purified DNA and culture material. For quantification, the *pla* target was used. Primers and probes were used as described by Riehm et. al.(12) for *pla*-specific qPCR and were obtained from TIB MOLBIOL (Berlin, Germany). The 20 µl ddPCR reaction mixture per sample contained 10 µl 2X Supermix for Probes (no dUTP), 1.8 µl of each primer (10 µM µl^-1^), 0.5 µl probe (10 µM µl^-1^) and 2 µl template. Droplets were generated using the BioRad QX100 Droplet Generator Hercules, USA) and PCR was performed on the Veriti 96-well Thermal cycler (Applied Biosystems, Foster City, USA) using the following reaction conditions: 10 min at 95 °C, followed by 40 cycles of 30 s at 95 °C and 1 min at 60 °C and a slope of 2 °C s^-1^. For enzyme deactivation, a final heating step of 10 min at 98 °C was performed at the end. ddPCR was evaluated using the BioRad QX100 Droplet Reader (Hercules, USA) and the corresponding QuantaSoft Analysis Pro software.

For precise calculation of number of cells per mL, absolute copy numbers determined by ddPCR were divided by 186, as *Y. pestis* hosts 186 copy numbers of the plasmid (pPst/pPCP1) encoding *pla* (13).

## RESULTS

To assess overall functionality and applicability, we applied the PCA method to detect mecA. As template, we used purified genomic DNA of MRSA, which was denatured at 99 °C for 10 minutes (1 min was also shown to be sufficient, data not shown). With this assay, down to 10 copies per reaction could be detected reliably, illustrating the sensitivity potential of PCA. However, the onset of amplification did not clearly depend linearly on the logarithm of the initial copy-numbers, indicating that PCA in its current implementation is a qualitative or only semi-quantitative method (Fig. 3)

**Fig 3:**
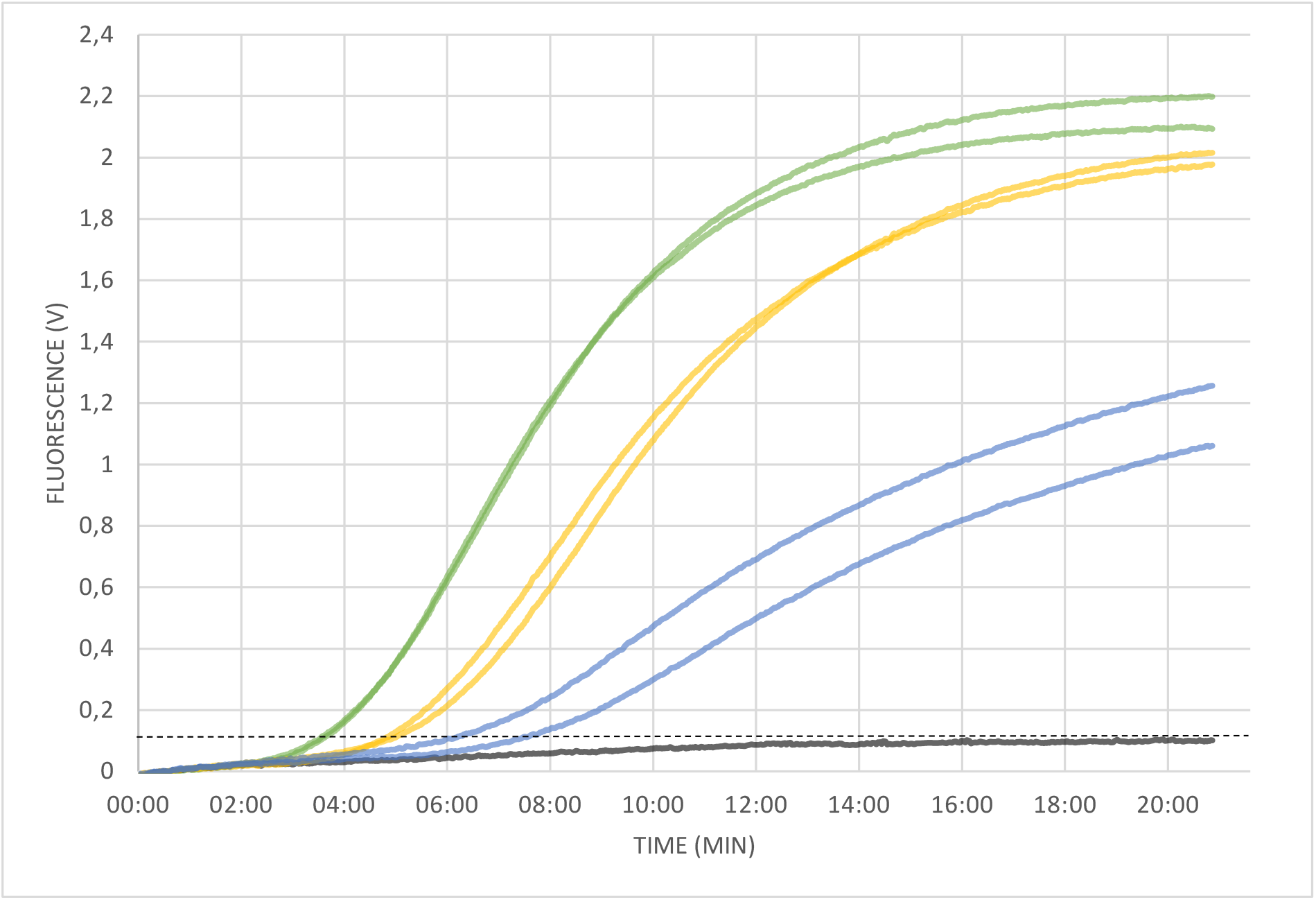
Detection of MRSA mecA gene. PCA using primers specific for *S. aureus* mecA gene with purified and denaturated DNA as template. Different DNA concentrations were used in duplicate reactions. Exponential amplification was observed for all template containing samples: 10^3^ (green), 10^2^ (yellow), 10^1^ (blue) copies per reaction. No amplification was observed for the negative control (black)

Subsequently, we applied PCA to *Y. pestis*, using the pla gene as target and purified genomic DNA as template. While the negative control did not produce a signal, clean amplification was observed in all template-containing samples (in duplicates). The detection threshold was reached in 03:54 minutes (10e8 copies) and 6:50 minutes (10e5 copies) and the entire PCA run was completed in 13.75 minutes. Again, the onset of amplification did not clearly depend linearly on the logarithm of the initial copy-numbers (Fig. 4A).

**Fig 4:**
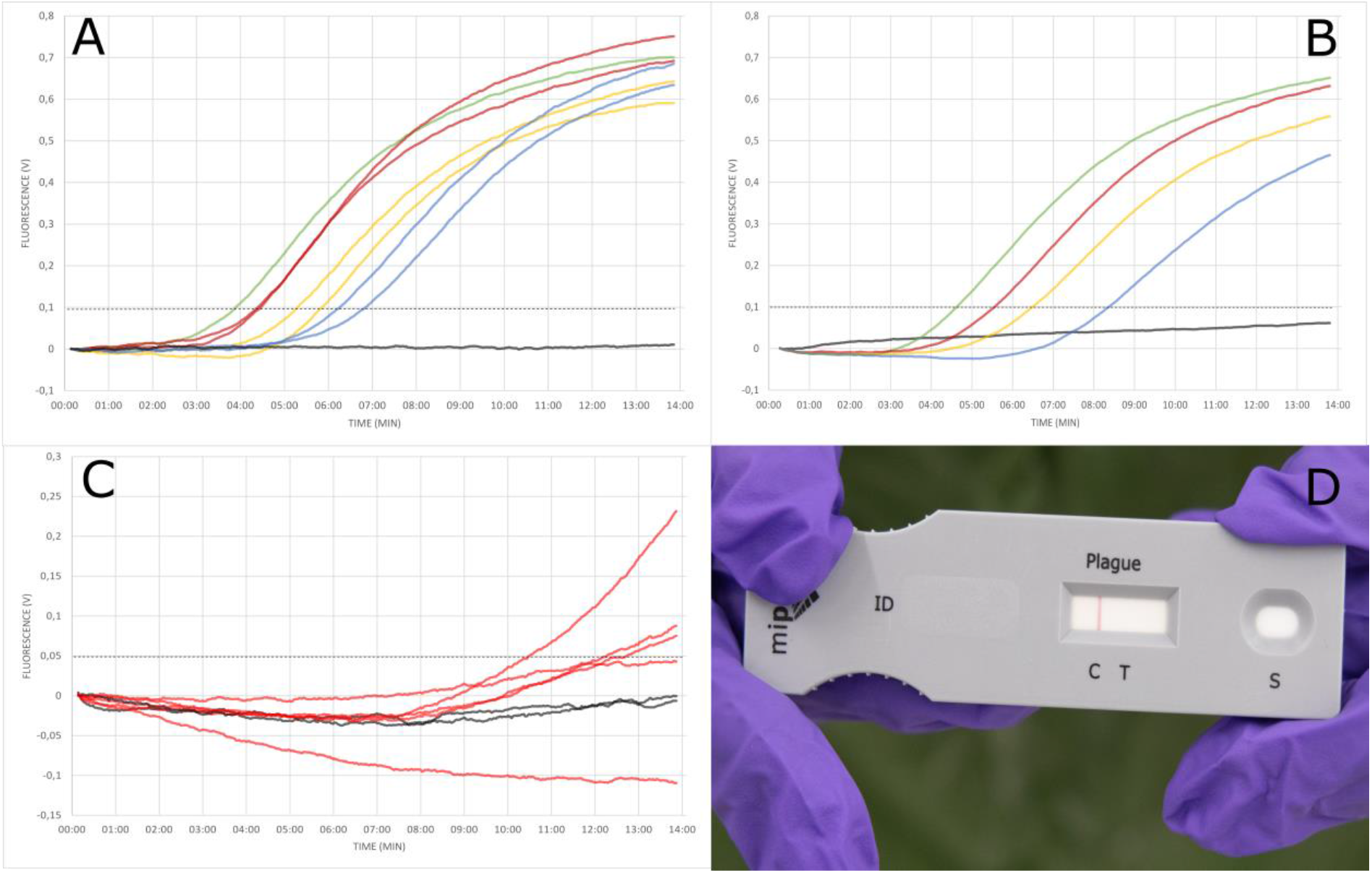
Detection of *Y. pestis* pla gene. **(A)** Different DNA concentrations were tested in duplicate reactions using purified DNA as template. Exponential amplification was observed for the positive control containing 10^8^ copies (green), and all template containing samples: 10^7^ (red), 10^6^ (orange), 10^5^ (blue) copies per reaction. No amplification was observed for the negative control (black). The onset of amplification did not clearly depend on the logarithm of the copy-numbers used, indicating a semi-quantitative quality of PCA **(B)** *Y. pestis* specific PCA was performed with crude sample material and an initial 5-minute hybridisation step prior to PCA. Exponential amplification was observed for positive control (green), undiluted sample (1.3 x 10^8^ cells per ml; red), 1:10 dilution (yellow) and 1:100 dilution (blue) of sample. No amplification was observed for sample containing no bacterial culture material (black). **(C&D)** PCA and LFA specific for *Y. pestis* were performed in parallel under field condition wearing heavy PPE. Results were obtained within 20 minutes for both tests. PCA: negative controls (black), sample (red). LFA: test was valid as indicated by the control line but was negative for *Y. pestis*

To assess field applicability, we applied the same PCA protocol to crude culture material of *Y. pestis* and added an additional 5-minute incubation step at 66 °C base temperature. Initial experiments were performed with performed with PFA-fixated sample material and clean amplification was observed for all tested log-dilutions of sample while negative controls did not produce any signal (data not shown). Subsequently, all experiments were repeated with 10-fold serial dilutions of non-fixated sample material and observed the same results (Fig. 4B)

### Determination of precision and limit of detection of the Y. pestis pla assay

Copy numbers of serial dilutions of both purified DNA and culture material were determined by ddPCR. The sensitivity and reproducibility of PCA were then investigated using probit analyses. Each PCA was performed three times with six replicates for each tested dilution. Results revealed a LOD (95%) of 434 copies per reaction for purified DNA and 9.8 cells per reaction (≈164 cells/mL), respectively, and indicated efficient reproducibility of the assay. Detection of lower copy numbers was possible but less reproducible for the pla PCA assay.

### Evaluation of specificity

To determine specificity of the pla assay, PCA was performed in duplicates with purified DNA of various bacteria listed in table S1. Based on whole genome sequences, *Y. enterocolitica* and *Y. pseudotuberculosis* are classified as nearest phylogenetic neighbors (14). For these closely related species, specificity was tested in triplicates with not only purified DNA but also bacterial culture material. For all bacteria tested, PCA was negative, confirming the specificity of the assay.

### Evaluation of field handling

To evaluate feasibility of the method under field conditions, both PCA and LFA were performed in parallel with operators equipped with heavy personal protective equipment (PPE) in a training scenario during the international CBRN-live agent exercise Precise Response 2019 in Suffield, Canada. In this scenario a clandestine laboratory was searched by special forces wearing fully encapsulated PPE. A turbid liquid broth incubated at 28C° and scientific literature evidence pointed towards an attempt to grow *Y. pestis*. After sampling of the liquid bacterial culture, both tests were performed simultaneously. Results were obtained after 20 minutes. While the LFA was unable to detect *Y. pestis* (Fig. 4D), PCA was positive in three out of five replicates, while both negative controls did not produce a signal (Fig. 4C). The sample was then transported back to the stationary lab and DNA was extracted. Subsequently performed qPCR confirmed presence of *Y. pestis* in the acquired sample material along with large amounts of *Escherichia coli* (data not shown).

## Discussion

PCA is a novel method based on the principle of PCR, which can effectively be used in the detection of infectious agents. To compare PCA and LFA directly, an assay for the detection of *Y. pestis* was developed. Since PCA is based on nucleic acid amplification and detection *Y. pestis* pla gene was used as target. Because of its high copy number (186) among the Orientalis biovar, *pla* is the most suitable and commonly used target (12,13). The results in this study show that PCA can be applied to both purified and crude sample material with a quick workflow of approximately 20 minutes for crude sample material, which is comparable to the time required for most LFAs. The negative result of the simultaneously applied LFA can be attributed to the combination of (i) a target cell number below the detection limit and (ii) the fact that the cells were grown at 28°C, leading to reduced target antigen expression(15). For the field handling tests we challenged the prototype device by mixing the target bacteria with a large amount of non-target bacteria (*E. coli*), thereby simulating a contaminated culture in the clandestine laboratory. PCA was capable of detecting the target organism even within this massive non-target background. Even though we operated the device in the field under challenging conditions and at the limit of detection, the fact that 3 out of 5 replicates were positive illustrates the advantages over LFA and PCR of this technology and its potential for field applications.

The LOD (95%) of purified DNA was 434 copies per reaction, however since LFAs are antigen-based tests, results cannot be compared directly. As an indirect comparison, we determined that PCA is more sensitive in detecting *Y. pestis* in culture material with LOD (95%) of 164 cells/mL, which is a result of the high gene dose effect. This LOD is well below the described LOD for *Y. pestis* LFA – as illustrated by the negative LFA, which we simultaneously conducted in the field. When compared to *pla*-specific qPCR on the other hand, it becomes clear, that qPCR is more sensitive with published LODs (95%) of as low as 0.1 genome equivalents (16). However, as illustrated by the mecA assay, which displays comparable sensitivity to conventional real-time PCR, PCA sensitivity is assay-dependent. When comparing time to result, PCA is much faster in obtaining results (13.75 min total run time) than Real-time PCR (> 60 minutes). Taken together, the above results demonstrate that as an molecular diagnostic technology, PCA combines some of the most advantageous elements of both LFA and PCR: (I) it is more sensitive and specific than LFAs (II) it is as specific as PCR, (III) it takes a fraction of the time needed for PCR (IV) in contrast to PCR can run on a small, battery powered, portable platform that enables testing in extra-laboratory or non-traditional laboratory environments as well as the Point of Care (Tab. 4)

Due to its speed and portability, PCA facilitates DNA-based detection of infectious agents in a non-laboratory POC environment. In 2003, the WHO identified seven ideal characteristics of tests that all newly developed POC diagnostics should meet. Based on the acronym ASSURED, these tests should be **A**ffordable, **S**ensitive, **S**pecific, **U**ser-friendly, **R**apid & robust, **E**quipment-free and **D**elivered (17). However, even in 2020, hardly any tests exist that meet these criteria. While LFAs meet some of the criteria outlined by WHO, they greatly lack sensitivity and specificity as previously described. PCR on the other hand is neither rapid, nor equipment-free. In addition, it requires expensive instrumentation and trained medical staff to be performed and evaluated. PCA is rapid, sensitive, specific, and it requires only minimal equipment and is relatively simple to perform. However, PCA does require initial training to perform and evaluate an assay.

Due to the simplicity of PCA assay design, which requires the same number of primers as conventional PCR, it may be possible to transfer the majority of existing real-time PCR assays to the PCA format with little or no modifications/optimizations. Moreover, in conventional PCR, nucleic acid extraction methods are often necessary prior to amplification and detection – thereby prolonging the whole procedure and limiting its’ field applicability. PCA technology is also suitable for the multiplex format (data not shown). While other commercially available platforms, such as the film array (18) allow for highly efficient multiplex screening for several pathogens, their power requirements require they remain lab-bound and are not true point-of care or incidence technologies. In addition, the high instrument and assay costs further prevent widespread use of such devices. Finally, the PCA method allows for swift responses to emerging biological threats with a rapid on-site test by straightforward design of PCA assays based on laboratory PCR primers.

In conclusion, we demonstrated a novel ultra-fast method for the amplification and detection of nucleic acid though Pulse Controlled Amplification (PCA). PCA combines the speed of LFAs or isothermal methods, with the simplicity of assay design of PCR, in a novel portable system. This work successfully demonstrates example applications for PCA in the detection of MRSA and of *Y. pestis*, both lethal pathogens for which limited Point of Care diagnostic options exist, under both laboratory and field conditions. It expands the use of molecular testing to extra-laboratory or non-traditional laboratory settings, as well as near-patient setting, and has the potential to become a gold standard technology in nucleic acid detection for front-line and in-field applications.

## Data Availability

All data are fully available

## SUPPLEMENTARY

Technical information on the Pharos micro prototype: The thickness of the heating layer scales with 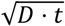, where D is the thermal diffusivity of the reaction solution (on the order of. 1.5. 10^−7^*m*^2^/*s*) and t being the heating duration. By this approach, the heated mass of the wires and the surrounding liquid within the heating layer is a minute fraction (≪ 1%) of the entire reaction volume. For the denaturation step, a 10 mF capacitator is loaded to 30-40 V, which is delivered for a sub-millisecond duration via a MOSFET (metal-oxide-semiconductor field-effect transistor; serving as a fast switch) to the wires that are heated to >90 °C to denature the amplicons on their surface. In order to keep the thickness of the heating layer as small as possible, a short heating duration is required, which can be achieved by applying a pulse at substantial peak power in the order of 1 kW to the wire array for a few hundred microseconds. Due to the short duration of each pulse, however, the average power consumption for the entire instrument is below 8 W (including the heat blocks, the fluorescence unit, the microcontroller and the pulsing).

**Tab S1.**
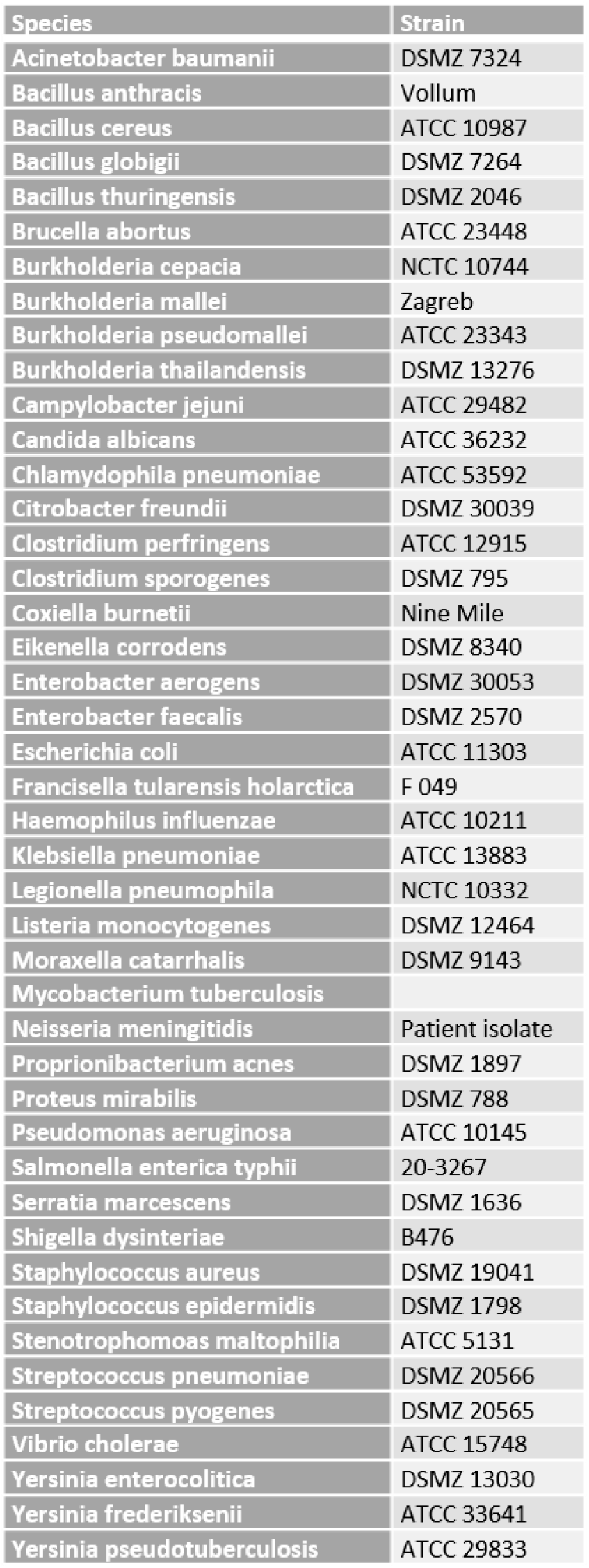
No cross-reactivity was observed with any of the tested bacteria, confirming specificity of the pla assay. (ATCC: American Type Culture Collection; DSMZ German Collection of Microorganisms and Cultures; NCTC: National Collection of Type cultures)

